# Global Patterns in Access and Benefit-Sharing: A Comprehensive Review of National Policies

**DOI:** 10.1101/2024.07.12.24310347

**Authors:** Gunnar V. Ljungqvist, Ciara M. Weets, Tess Stevens, Hailey Robertson, Ryan Zimmerman, Ellie Graeden, Rebecca Katz

## Abstract

**Introduction:** The goal of Access and Benefit-Sharing (ABS) in global health governance is to ensure that countries that provide genetic resources, including pathogens, receive equitable access to the benefits derived from their use. The increasing availability of genetic resource digitalization has brought this issue to the forefront of discussions on global health security and health equity. While originally conceptualized in supranational agreements, implementation of these treaties requires national-level legislation in each country. This work represents the first comprehensive effort to map ABS policies in all 193 United Nations member states.

**Methods:** We conducted a standardized review of the legislation for 193 United Nations Member States across 3 global legal databases (ABS Clearing House, WIPOLEX, and FAOLEX), national legal databases, and a systematic Google search. Legally-enforceable policies were identified, and data was extracted across the following 8 aspects of ABS legislation: Scope of Legislation, Digital Sequence Information, Access to Resources, Prior Informed Consent, Contractual Terms, Benefit-Sharing, Compliance, and Legal Sanctions.

**Results:** We found that 104 countries have legally-enforceable policies on ABS, with 92 countries having ABS policies relevant to microorganisms. Of these, 74 countries have chosen to restrict access to their domestic pathogens, and 53 have chosen to link access to pathogenic resources with an obligation to share benefits. Altogether 60 countries have a codified position on Digital Sequence Information (DSI) with regard to ABS: 20 have included it, 34 have excluded it, and 6 have ambiguous wording. WHO regional coverage of ABS or DSI policy ranged from 28% (3/11) of countries in the Eastern Mediterranean Region, to 62% (33/54) of countries in the European Region.

**Conclusion:** These findings highlight the heterogeneity found in the global policy landscape as it pertains to ABS, and provide data to inform future agreements and research efforts related to ABS.

**Key Questions:**

- Recent pandemics and technological advances have put Access and Benefit-Sharing (ABS) in the center stage of global health diplomacy. Yet, efforts to harmonize these policies have stagnated in multilateral negotiations. There is a distinct scarcity of evidence on the differing interpretations of ABS around the world, and further research is urgently needed to inform ongoing negotiations.
- This study provides the first detailed global mapping exercise of the ABS policy landscape. We found that while over half of the world’s countries have legally-enforceable policies relevant to ABS, only about a fourth have defined a position on Digital Sequence Information. There was also significant geographic variation in policy coverage within WHO regions.
- This study provides data to inform future research endeavors, highlighting global trends in national policy and identifying governance gaps. This open-source policy database could inform future evidence-based policy-making on ABS at the national level and enhance understanding of the current legal environment for ongoing negotiations on a Pathogen Access and Benefit-Sharing mechanism.

## 1. INTRODUCTION

Access and Benefit-Sharing (ABS) in global health security is the concept of controlling access to genetic resources to ensure the equitable distribution of benefits derived from their uses. ABS has become one of the most important, and at times divisive [1], issues in global health governance, and has emerged as a key factor in discussions around equity, information sharing, and outbreak response. The Convention on Biological Diversity (CBD), which entered into force in 1993, established ABS as a critical component of both the sustainable use of natural resources and conservation of biodiversity. Article 3 established the states’ sovereign right to control the use of genetic resources physically located within their borders [2]. CBD also affirmed that access to genetic resources was subject to national legislation, required prior informed consent (PIC), could be denied, and had to be granted to international parties on mutually agreed terms (MATs) [3]. In particular, it established that the benefits derived from access should be given, in priority and on a fair and equitable basis, to low- and middle-income countries (LMICs). In generating a new revenue stream for biodiversity-rich countries, the Convention endeavored to promote economic growth in LMICs while simultaneously creating financial incentives for governments to combat biodiversity losses due to deforestation, pollution, and biopiracy [4–6]. ABS mechanisms thus sit at the nexus of ecological legislation, economic policy, and global equity.

#### Box 1: Key Definitions

**Genetic Resources**: Usually taken to mean the physical samples derived from biological sources. In the context of pathogen research, this may refer to a pathogen sample, and by extension any medium they may be suspended in or grown on.

**Biological Resources:** a broader term than “genetic resources”, often encompassing plants, animals, and microorganisms, including their genetic components as well as non-genetic derivatives. Sometimes used interchangeably with “flora & fauna” or “natural resources”. These terms are usually defined explicitly in individual policies, and the exact extent of their scope can vary.

**Digital Sequence Information (DSI)**: This refers to the description of genetic resources in terms of its constitutive genetic nucleotide base pair sequences, rather than genetic material. While the use of the term DSI over the term “Genetic Sequence Data” (GSD) is a matter of some debate, in this paper these terms are used synonymously for the sake of clarity.

To support implementation of the principles outlined in the CBD, the Nagoya Protocol on Access to Genetic Resources was adopted in 2010 [7]. This protocol, open to Member States of CBD, further developed the language to safeguard traditionally exploited entities, including indigenous groups, to ensure equitable negotiations between providers and users of genetic materials [8,9]. Taken together, these agreements, rooted in biodiversity conservation efforts, provided a foundational legal framework for the transfer of biological materials.

While originally conceptualized in biodiversity agreements, ABS was recognized to have implications for pandemic preparedness and response, particularly with respect to pathogen sharing. In 2007, during an outbreak of H5N1 influenza in Indonesia, the Minister of Health announced a decision to cease pathogen sample sharing with the World Health Organization (WHO) and other global partners after a company in a High-Income Country (HIC) expressed interest in creating a vaccine from Indonesian samples. Indonesia cited CBD, claiming that a country’s right to control its genetic resources explicitly included those pathogens isolated within its borders, and used the principle of “viral sovereignty” to force discussion of more equitable access to benefits (e.g., vaccines) derived from free sharing of such samples [10–12]. However, the exertion of such sovereignty was controversial, as timely sharing of pathogens with pandemic potential within the international community facilitates surveillance activities and the development of medical countermeasures [13]. Thus there were concerns that sovereignty claims would delay timely sharing of pathogens [14,15]. The debates around “viral sovereignty” influenced the Nagoya Protocol negotiations and also led to the development of a new agreement under the auspices of the WHO. Adopted in 2011, the Pandemic Influenza Preparedness Framework (PIP) was the first international agreement to acknowledge viral sovereignty and address the need for a mechanism that could rapidly share pathogen materials while simultaneously upholding the principles of ABS [16]. Within PIP, this mechanism took the form of legally-binding terms using Standard Material Transfer Agreements (SMTAs) that would be applied to all entities internationally transferring influenza genetic resources. The intent was that the SMTAs would ensure an equitable process for sharing and deriving benefits from influenza genetic material, including access to vaccines in the case of an outbreak.

A robust ABS system has become increasingly critical to many countries, particularly LMICs. Due to the growing use of genomic sequencing and DNA resynthesis technologies, information regarding genetic resources has become an important commodity [17]. As these technologies usher in a new era of pathogen data sharing, in addition to concerns over sharing physical samples, there is need to consider Genomic Sequence Data (GSD), or Digital Sequence Information (DSI), as it is referred to within the CBD community. During the 2015 outbreak of Ebola in West Africa, DSI was used by entities from HICs to develop a novel medical countermeasure from genomic sequence data isolated in Guinea and publicly shared [18]. As this product was developed based upon freely available DSI, there was no explicit obligation under the existing ABS framework for the developer to share benefits, monetary or non-monetary, with the country of origin. But the question of whether DSI is already captured by ABS policies under the definition of “genetic resources” is a controversial one [19]. This issue was further underscored during the COVID-19 pandemic, where millions of genetic sequences were shared, highlighting the differences in the platforms used to share this information, and the disconnect between sharing DSI and eventual access to medical countermeasures [20–22].

Catalyzed by the inequities identified during the COVID-19 pandemic, the CBD Secretariat declared in 2022 the intention to implement a unifying ABS mechanism for DSI to generate appropriate benefits for donor countries while ensuring legal clarity and efficient access [23]. Similarly, negotiations for a new international agreement on pandemics include a proposal for a new Pathogen ABS (PABS) system [24].

At the 77^th^ World Health Assembly in 2024, member states were unable to agree on this new international agreement and were granted a one-year extension for negotiations [25]. The failure to reach an agreement in 2024 highlighted the ideological divisions, geopolitical power dynamics, and diverging interests that render supranational negotiations on ABS challenging [26]. National governments, however, have long employed country-level policies to govern the transfer of genetic materials, and sometimes, explicitly DSI. In fact, parties to Nagoya are obliged to develop implementing legislation under Article 8 [7]. Expanding upon previous case-study research on specific aspects of ABS policy [27,28], our research systematically maps and analyzes the national-level policies from the 193 UN Member States on ABS as they relate to microorganisms. To our knowledge, this is the most comprehensive mapping of national-level policies governing all aspects of ABS for both pathogens and GSD. We believe this mapping to be a critical foundation for understanding the state of ABS in the world, and critical to future bilateral, regional, and global negotiations.

## 2. METHODS

### 2.1 Project scoping and country inclusion

We identified and analyzed national policies relevant to ABS and DSI for all 193 UN Member States, regardless of their membership in international agreements, including the Nagoya Protocol, in order to provide an accurate and up-to-date reporting of the global policy environment (Figure 1).

**Figure 1:**
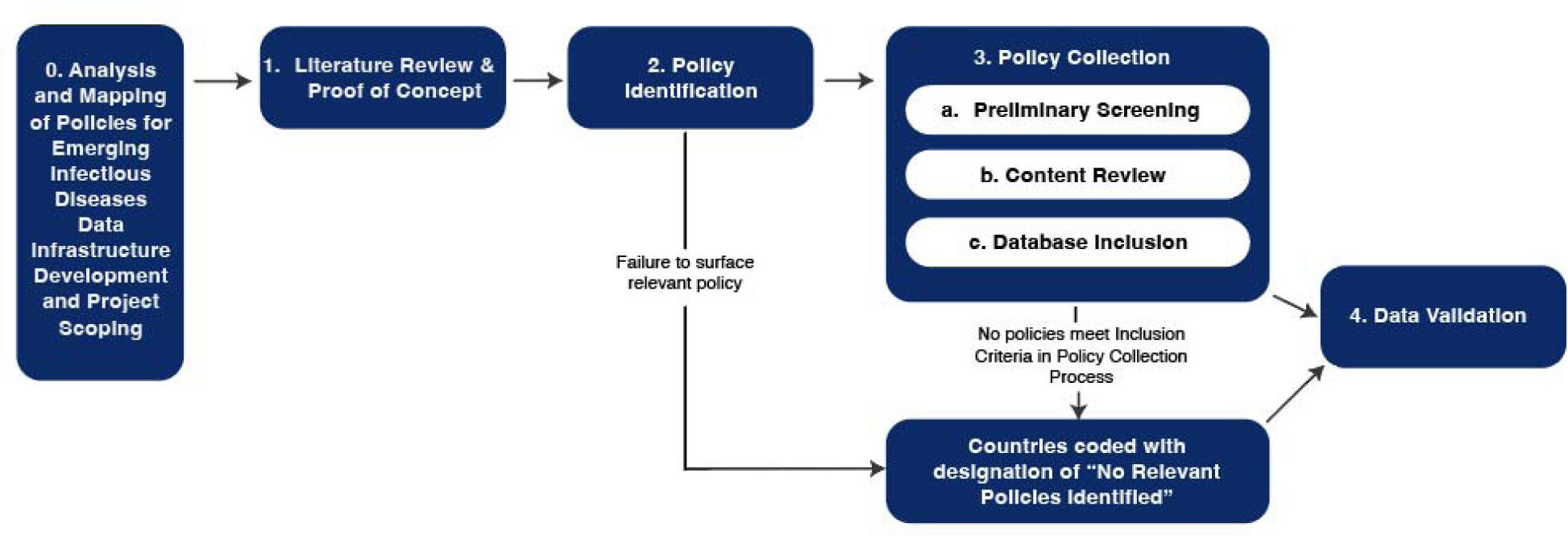
Methodology protocol utilized in the Analysis and Mapping of Policies for Emerging Infectious Diseases topic on Access and Benefit-Sharing. Numbering illustrates the sequential approach to project completion.

This ABS dataset is just one aspect of the multifaceted Analysis and Mapping of Policies for Emerging Infectious Diseases (AMP-EID) research effort, which seeks to create an unparalleled database of global outbreak preparedness and response policies across various topics. We employ a standardized operating procedure (SOP) across all policies for data collection. The SOP includes a literature review for each topic to identify relevant search terms for the topic’s policy collection protocol. After the completion of the ABS literature review, we conducted a proof-of-concept study of ten geopolitically and economically diverse countries to test the methodology and determine a final series of query terms to be employed. After proof-of-concept, we reviewed and resolved gaps in the policy collection protocol and coding methodology. The research team then created the customized data taxonomy (Supplementary Figure 1).

### 2.2 Identification of relevant policies

To collate a comprehensive dataset of relevant policies, we developed a standardized, sequential policy identification protocol for each country. Initially, three disparate online databases containing potentially relevant policies were systematically investigated. The ABS Clearing House was first consulted [29]. This online platform, administered by the Convention on Biological Diversity Secretariat, allows countries to upload their relevant ABS legislation thus facilitating sharing of information between signatory nations. We reviewed each country’s profile and downloaded all available active legislation for review. We then utilized the World Intellectual Property Organization legal database (WIPOLEX) [30]. This filterable database contains legislation relevant to intellectual property in each country and is centrally administered by the WIPO. We applied a filter for the subject matter of “Genetic Resources” to the legislation included on the platform for each country. If such policies were available, they were downloaded. Finally, we examined the Food and Agriculture Organization’s legal database (FAOLEX), one of the world’s largest legal databases [31]. We manually reviewed the Environment and Wild Species and Ecosystems categories of policies for each country. Laws directly mentioning “Access and Benefit-Sharing”, and “Genetic Resources” were all included.

Relevant overarching acts containing the terms “Biological Diversity/Biodiversity”, “Nature”, “Natural Resources”, “Environmental”, “Conservation”, were also included, unless explicitly unrelated (for instance, the Japanese “Act preventing the Environmental Pollution of Mercury”). We reviewed and screened the full text of these overarching acts for relevant sections to ABS, and if they lacked any mention of “access and benefit-sharing” or “genetic/biological resources”, they were subsequently excluded. All remaining legislation was deemed relevant and included for further analysis.

After completion of database consultation, we conducted a manual search of the national government’s legal database, where possible. If a legal database maintained on a country domain was available and open-access, all potentially relevant policies, utilizing the collection criteria described above, were captured.

To ensure comprehensive inclusion of potentially relevant policies, we then completed a manual search in the Google search engine using a standardized series of search terms developed through the literature and landscape review (Supplementary Table 1). For countries that conduct government in a language other than English, a machine translator was used to translate the series of search terms into the language primarily used by the government in the target country.

Once all potentially relevant policies from a country were surfaced, we used Google Translate to complete translation of policies in languages not spoken by the research team. Where possible, fluent speakers of non-English languages were contacted to verify machine translations.

In the case that this standardized collection protocol failed to surface any potentially relevant policies, we coded the country as “No ABS legislation identified”. Importantly, we identified all relevant, legally-enforceable policies in a country, though research on the extent to which any of these policies are implemented and enforced in the country was beyond the scope of this project. Thus, it is possible that execution of policies may be inconsistent with the policy mandate.

All policy identification and collection took place between October 2023 and May 2024.

### 2.3 Database creation

#### Primary and Secondary Policy Review by Inclusion Criteria

Potentially relevant policies identified through the standardized collection process were subject to a preliminary screening to eliminate documents that were not legally binding or were no longer enforceable. Thus, strategies, reports, and draft laws, as well as laws that had been repealed, were excluded. As an exception, supporting documents designed to clarify the interpretation of associated active laws or policies were included to provide critical contextual information. These entries have been clearly identified as supporting documents in the dataset.

Policies that passed the preliminary screening stage were then reviewed by the standardized inclusion criteria (Supplementary Table 2). Policies that met the inclusion criteria were categorized into the customized data taxonomy and countries were assigned an applicable status for each subtopic (Supplementary Figure 1). Policies were then downloaded as PDFs and collated in Airtable, a cloud-based platform for relational databases. All documents included in this dataset are publicly available for download in a comma-separated values (.csv) file.

#### Data Validation

Literature review, collection protocol, and inclusion criteria were reviewed by the entire research team and approved by the Principal Investigator. Policy collection and primary review by inclusion criteria were completed by a lead researcher. Once included in the database, a second member of the research team completed a secondary review of policies, assessing the primary researcher’s coding. Any coding discrepancies that arose between researchers were deconflicted and reviewed by the Principal Investigator. Finally, the data went through a quality control review by the research team before being uploaded to ampeid.org.

#### Data Availability

All data are available in a public repository at ampeid.org. All datasets, reproducible code, and figures are available at https://github.com/cghss/ABS [32].

## 3. RESULTS

We found ABS policies were applied, either explicitly or implicitly, to a broad range of sectors from fisheries to agriculture. Of the UN member states reviewed, 54% (104/193) had broad legislation pertaining to ABS in any context, although 12 countries, such as Mongolia or Niger, had legislation that did not apply to microorganisms, referring for instance to “plants and animals” [33,34]. Across these 104 countries, 181 total documents containing ABS policies were identified. Therefore, some countries employed multiple policies to regulate ABS, while others use a single policy to do so. In comparing our results with the existing database of ABS-Clearing House overseen by the CBD, we found that, of the 181 documents our methodology identified, 61% were also found in the ABS-Clearing House (111/181). Just under half of countries, 46% (89/193), have not codified legally-enforceable policy on either ABS or DSI.

Relatively few countries (31%; 60/193) had included language referring to DSI in enforceable policy or supportive documents. In many cases, DSI and ABS are addressed in the same policy, however this was not universally true, meaning that some countries had either ABS legislation that did not include provisions on DSI or standalone positions on DSI but no ABS legislation. For example, the United States and Canada, both members of the WHO Region of the Americas, were not found to have enforceable ABS policy at the national level, yet do have codified positions on DSI [35]. Conversely, San Marino and North Macedonia, both members of the European Region, have policies addressing ABS, but not DSI. This lack of uniformity accounts for discrepancies between the total number of states within a WHO region with ABS or DSI-related policies and the number of states within the region with policies that pertained to individual policy categorizations (Figure 2).

**Figure 2:**
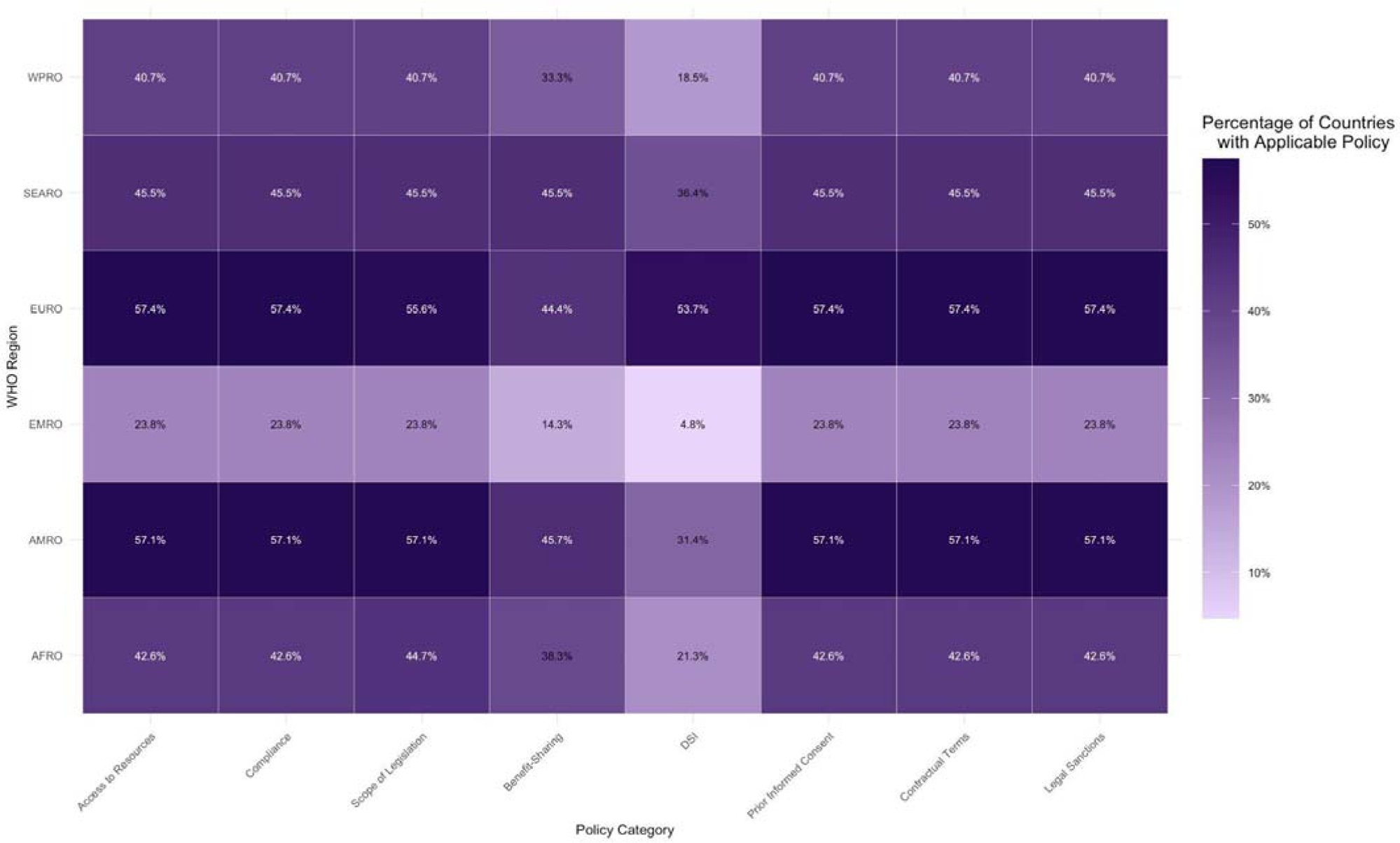
Heat map of percentage of countries within World Health Organization regions with legally-enforceable policy applicable to each subtopic. Shading represents the percentage of countries with applicable legislation, with darker shading indicating a higher percentage of countries with identifiable legislation pertaining to that subtopic category. Black numbering represents percentages less than 40.0%, while white numbering demonstrates percentages over 40.0%. Percentages were calculated as the number of countries within a WHO region with applicable policies for a subtopic divided by the total number of countries in the WHO region and rounded to the nearest tenth.

### 3.1 Regional Variations in ABS and DSI Policies

There is substantial interregional variation in the presence of legally-enforceable ABS and DSI policy. No WHO region had greater than 62% of countries with applicable policies. However, some regions had a significantly higher proportion of countries with applicable legislation than others (Figure 2).

The European Region and Region of the Americas had the greatest ABS and DSI policy coverage with 62% (33/54) and 60% (21/35), respectively. In the European Region, 57% (31/54) of countries were found to have policies addressing ABS, and 53.7% of countries address DSI. Importantly, these numbers are bolstered by a DSI law applicable to all European Union (EU) member states. Italy, Slovenia, and Lithuania have not yet codified this EU legislation at the national level, but were coded as having addressed DSI due to their mandate to do so under supranational law. The Region of the Americas has a similar gap between the percentage of countries that have ABS-related legislation (60%), and those with a policy that addresses DSI (31.4%). While both regions have a majority of states with ABS-related policies, policy coverage related to DSI remains less universal.

Nearly half of the states in the African and Western Pacific Regions have legislation pertaining to ABS or DSI. In the African region, 51% of countries (24/47) have legislation applicable to either designation, with 51% (24/47) of countries identified with ABS-related policy and only 21.3% (10/47) of countries identified with policy pertinent to DSI. 46% of countries in the Western Pacific Region were found to have DSI- or ABS-related policies. In this region, 41% (11/27) had policies that met inclusion criteria for ABS designation, while 18.5% (5/27) of countries had addressed DSI.

The Southeast Asian and Eastern Mediterranean Regions were found to have the lowest proportion of states with policies with ABS or DSI national-level policies. Of the Southeast Asian countries, a total of 45% (5/11) had policies related to ABS and 36.3% (4/11) had policies related to DSI. This region had the second-lowest proportion of states with policies pertaining to ABS, yet had the second-highest percentage of countries with DSI legislation. The Eastern Mediterranean Region had the lowest total policy coverage. Just 24% (5/21) of countries in this region were identified as having either ABS- or DSI-related policies, though only 5% (1/21) were identified as having a policy related to DSI.

### 3.2 Diversity in Policy Content

#### Access Restrictions and Scope of Application

Policies governing the access of international entities to domestic microorganisms were identified in 48% of countries (92/193). A further 46% (89/193) of countries were found not to have any national-level policies relevant to ABS, while the remaining 6% (12/193) were found to have legislation that could broadly be interpreted to encompass ABS but did not include provisions on microorganisms. Therefore, 52% (101/193) of studied countries were found not to have any national-level policies relevant to ABS applicable to pathogens.

Of those countries with relevant policies, 89% (82/92) encompass access to genetic resources, including microorganisms, as well as traditional knowledge, while the other 11% (10/92) only include genetic resources, including microorganisms. Of the 92 countries with policies regulating access to genetic resources, 80% (74/92) chose to restrict access. These restrictions may be stated explicitly, as in Algeria, which comprehensively outlines the assessments and declarations needed to garner access to resources [36], or implicitly, as in Tanzania, where resource sovereignty is codified, but regulatory mechanisms have not been established [37]. Access restrictions included in relevant policies may also differ based on the intended use of the extracted resource. For instance, as a precondition to access, Filipino law requires a Commercial Research Agreement if genetic resources are intended for commercialization, whereas an Academic Research Agreement suffices otherwise [38]. The remaining 20% (18/92) of countries with policy on resource access opted for unrestricted access. This may be done through explicit provisions in legislation, as is the case in Japan [39], or through implicit authorization. For example, many EU countries omit any access restrictions in relevant policy, thereby tacitly permitting extraction and usage of their genetic resources [40,41].

#### Preconditions for Resource Access

A subset of countries’ national-level policies require benefit-sharing, consisting of either monetary benefit-sharing such as royalties or milestone payments, or non-monetary benefits such as technology transfer and academic credentials, as a precondition for access. We found that 53 countries have legislation linking benefit-sharing with access. For instance, in Brazil, the various modalities of benefits required to gain access to genetic resources are outlined in relevant law [42]. The remaining 28% (21/74) do not explicitly establish an obligation to share benefits related to accessed genetic resources, despite having restricted access. This is the case for the Central African Republic, where the law states that the sharing of benefits should be “taken into account”, but is not explicitly mandated [43] (Figure 3).

**Figure 3:**
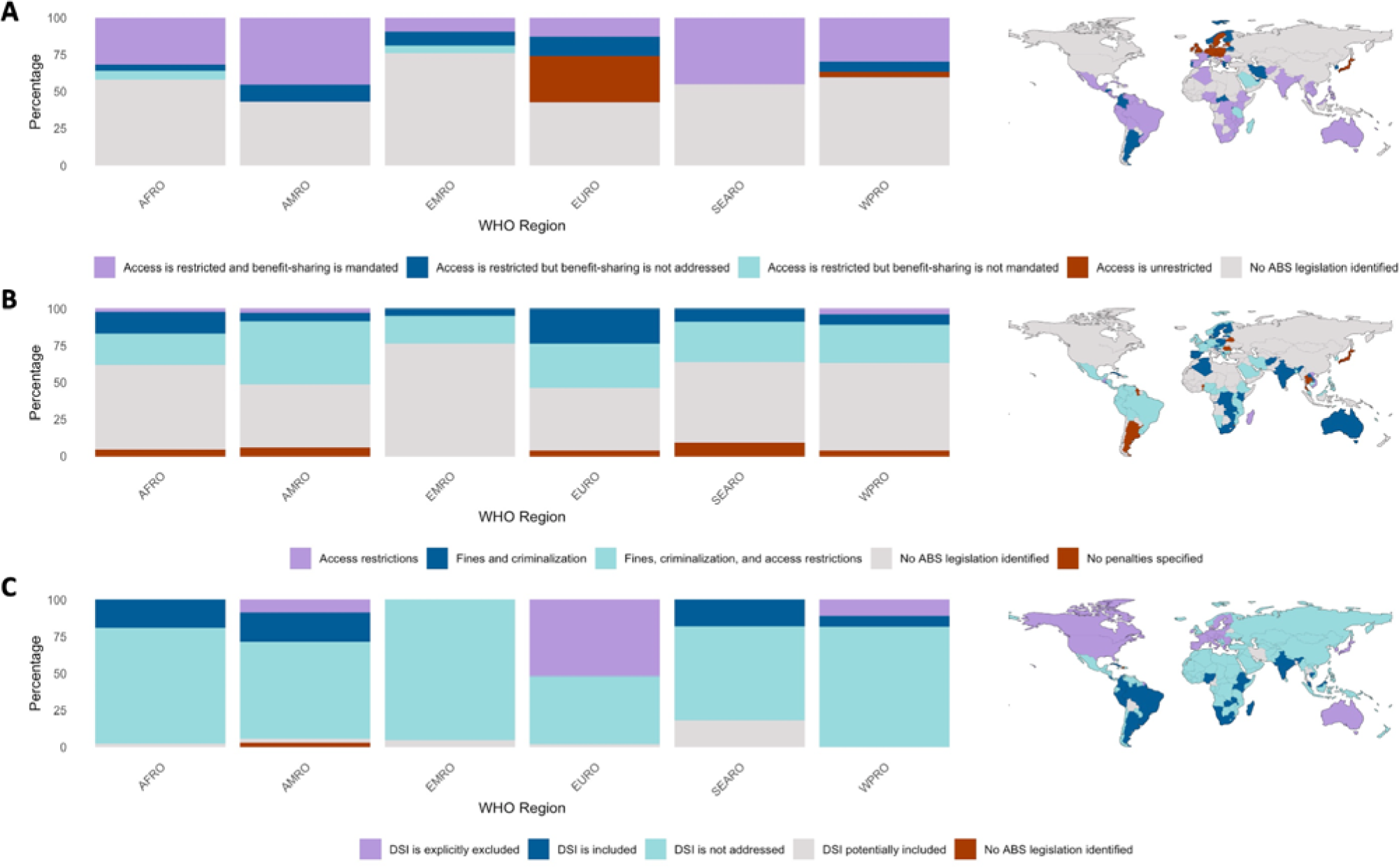
Graphical and mapped representation of countries with (A) legally-enforceable policy pertaining to genetic resource access, (B) sanctions included in legally-enforceable ABS policy, and (C) legally-enforceable policy that covers Digital Sequence Information (DSI). Bar graphs are arranged by WHO regions, with bars illustrating the percentage of countries within the region with identifiable policy for each of the subtopics. The corresponding maps, located to the right of each bar graph, demonstrate country-level status.

PIC is a consent mechanism that requires extractive entities to inform relevant authorities of the materials sought, their intended use, and the project timeframe as a precondition to resource access. The 20% (18/92) of countries that have codified open access to domestic genetic resources in policy, by definition do not require PIC agreements. However, of the 74 countries that do restrict access to resources, PIC mandates are codified by 82% (61/74) of the countries that restrict access to genetic resources. Often, these provisions have been included in legislation to protect indigenous populations, as is the case in Zimbabwe. Zimbabwean law outlines requirements for consent-seeking and encodes the rights of indigenous communities to be consulted in granting access to indigenous genetic resources for non-indigenous entities [44]. The remaining 18% (13/74) have not included PIC provisions in their legally binding ABS policies.

Separately, states may also require a written contract for ABS, which often takes the form of Material Transfer Agreements (MTAs) or Mutually Agreed Terms (MATs). Of the 74 countries that restrict access to resources, 74% (55/74) require the establishment of a contractual agreement between the resource provider and the extractive entity, while the remaining 26% (19/74) of countries have not established a requirement for contractual terms despite having legislation restricting access to resources.

#### Compliance Mechanisms

The vast majority of countries (95%; 87/92) that restrict access to genetic resources include provisions for compliance mechanisms in legally binding policy. However, countries use diverse approaches to enforcing their ABS laws. Malaysia, for instance, requires those international entities exporting genetic resources to pass through checkpoints [45], while France empowers specific enforcement officers responsible for ensuring compliance with ABS policies [46].

Legal sanctions, or punishments for international entities found to have violated ABS legislation, are less commonly included in policy than mechanisms to proactively ensure compliance. Of the countries that restrict access to genetic resources, 91% (84/92) include legal sanctions in their legally-enforceable policies (Figure 3). Such sanctions may include fines, criminalization, and further restrictions on access. Of these, some states (4%; 3/84) solely employ restrictions on access as sanctions, such as in legislation from Madagascar, which established the revocation of access permits and bans on future access [47]. Others (31%; 26/84) chose to use fines and criminal processes. More commonly these sanctions are used in concert, as is the case in Cambodia where delinquent entities are both charged a monetary fine and access permits are revoked [48]. The combination of fines and criminal prosecution with access restrictions was the most commonly identified legal sanction (65%; 55/84).

#### Digital Sequence Information

While not explicitly included in the CBD, Nagoya Protocol, or other international agreements, international sharing of DSI may be included in national-level ABS legislation or may be governed by an entirely distinct DSI policy. As of May 2024, 69% of countries (133/193) had not codified a position on DSI in legally-enforceable policy, leaving 60 countries (31%) with any legislation pertaining to DSI (Figure 3). Of these, 33% (20/60) of countries explicitly stated that DSI was included in their ABS legislation, while a further 57% (34/60) explicitly excluded DSI from ABS policies. A 2009 Nigerian policy, for instance, includes DSI by stating that access to genetic resources pertains to all “intangible components”, as defined by “information associated with or regarding genetic resources” [49]. In contrast, the Republic of Korea clearly articulates that genetic sequence data does not constitute a genetic resource [50]. The European Union, similarly, has asserted that in the absence of a global consensus on the term “DSI”, it would view it as distinct from “Genetic Resource” and thus not covered by ABS policy [51]. Some countries (10%; 6/60) included ambiguous language which may be interpreted to cover DSI, without explicitly doing so. For example, ABS legislation from Bangladesh includes, “biological material including (…) its various expressions and embodiments in knowledge”, which might effectively cover DSI [52].

## 4. DISCUSSION

Access and Benefit-Sharing policy is a critical part of the current governance between nations engaging in the international transfer of genetic resources and information. Mapping these policies creates transparency around processes in place to govern equitable relationships between entities in compliance with national-level policy, and facilitates access to information by providing all parties with clarity on the policy landscape in partner nations. We found that 54% of UN Member States have publicly available, legally-enforceable ABS policy at the national level, while 31% of UN Member States have included language on DSI in enforceable policies. Moreover, there is significant variability within most WHO regions in the presence of legally-enforceable ABS and DSI policies, with no region found to have greater than 62% of countries with applicable legislation. Despite this interregional heterogeneity, we did find notable thematic trends. Policy categories that were broader, such as those affirming sovereignty and regulating access to genetic resources, as well as those that outlined legal sanctions and mechanisms to ensure compliance, were identified in a greater proportion of countries. By contrast, we found that subtopics representing niche areas applicable only to ABS-specific policy, such as benefit-sharing requirements and regulation of DSI, were less likely to exist in the current national policy landscape.

Regional variation in policy coverage may be largely explained by geopolitical relationships and economic factors. Regions with relatively strong cultural, political, and historical ties, including Western Europe, Andean South America, and Sub-Saharan Africa, tend to carry similarities in their ABS legislation. We found that ABS policy was coordinated to the greatest extent between countries within formal supranational organizations, such as the EU [51,53] or Andean Community [54]. The majority of the countries within these customs unions had reached broad consensus over their views on ABS, which were reflected in national-level policy. While not always geographically clustered, we also found that countries in similar economic strata tended to adopt substantively similar stances on ABS. For example, wealthier nations tended to either leave access to resources unregulated at the national level, as was the case in the United States and Canada, or explicitly allow for free access to domestic genetic resources, as is permissible in many Western European nations and Japan. Conversely, many LMICs, particularly in areas known to be highly biodiverse [55], were more likely to have adopted national legislation restricting access to genetic resources, requiring benefit-sharing, and creating compliance mechanisms relative to their wealthier counterparts.

Variation in policy coverage by category was observed across regions, suggesting similar factors affected the inclusion of certain aspects of policy across geographic and economic groups. Legislation that broadly affirms state sovereignty over genetic resources and defines which resources fall under legal protection are the most commonly identified, but also the most diverse and unclear. Often these laws predate discourse on bioprospecting, yet the laws establishing resource sovereignty have been retrofitted to cover genetic resources or traditional knowledge, contributing to the observed heterogeneity and ambiguity of their contents [56]. For example, some policies restricting access to resources fail to explicitly define the scope of the protections, create unclear compliance processes, or simply fail to outline any pathway to resource access. Nevertheless, the vast majority of countries that restrict access to genetic resources include both preconditions to resource access, in the form of PIC and benefit-sharing, as well as compliance mechanisms and legal sanctions in legally-enforceable policy. However there were notable qualitative differences between countries in these categories that may have implications for equitable sharing of benefits derived from resources and compliance with these policies. Despite variations in how countries govern preconditions to access and mechanisms for policy enforcement, the commonality of these provisions from countries that restrict access to genetic resources reflects an interest from these nations in ensuring equitable access to the benefits derived from their resources and enforcing their ABS policies.

DSI sharing was the least covered policy category. When addressed, the terminology used was inconsistent, including “sequential information” [57,58], “genetic heritage” [35,42], “intangible components” [49,59–61], “information related to genetic resources” [45,62,63], and “utilization of resource” [35,64]. These inconsistencies in the policy environment suggest a lag time between advances in data-sharing techniques and the development of legally-enforceable policy. Indeed, the multilateral community has yet to agree upon a concrete definition of DSI, let alone tackle logistical issues such as storing, tracking, and intellectual property [65–68]. Considering the fast pace at which DSI is becoming the norm for pathogen sharing, any future attempts at reforming ABS policy are likely to be undermined unless they address these questions around DSI.

There are efforts within the international community to try to address both ABS policy heterogeneities and variation in how DSI is addressed. A Pathogen Access and Benefit-Sharing (PABS) system has been proposed as part of negotiations on a new Pandemic Agreement [24], with the intention to harmonize the various ABS processes and terminology across all member states. Additionally, the World Intellectual Property Organization (WIPO) has debated compliance mechanisms in international patenting laws of biotechnology, starting with the Colombian proposal of 1999 [69], and settling in 2024 on a treaty establishing a disclosure requirement when applying for a patent based on a genetic resource [70]. As negotiations resume ahead of the 78^th^ World Health Assembly, these issues are set to be at the center of discussions moving forward.

### Limitations

This research has important limitations and creates opportunities for further study. The policy identification protocol relies on the digitization of policy in publicly available repositories. Thus, nations without freely accessible online policy databases may not have been accurately captured in this work. Additionally, this study was constrained to national-level policy, though some countries regulate ABS or DSI at the subnational level. For example, Australia utilizes a combination of subnational and national level policy to regulate both ABS and DSI [27], however, in accordance with the project methodology, we captured only the policy enforceable at a national level. Given the complexity of this issue, for countries that utilize subnational policy to govern ABS, a case-study approach may be more appropriate to accurately capture the policy environment. Furthermore, we were only able to analyze what was explicit in the policy, though implementation and enforcement of these policies may be inconsistent with the policy mandate. Without data on the implementation and enforcement of these policies, including local cultural contexts, our results should only be considered informative of the current policy landscape.

## 5. CONCLUSION

ABS is a critical part of global health governance, and, from past events, one of the most controversial. The world, however, will continue to experience emerging infectious diseases and pandemics, technology will continue to trend to digitalization, and nations will continue to exert their sovereignty in service of national interests.

As nations negotiate ABS in a multilateral setting, it is critical to understand what policy already exists and where differences emerge. This research supports engagement by both researchers and other governments in approaching their own ABS legislation. It also provides an opportunity for empirical research and examination of national-level ABS legislation for specific outcomes. Such efforts will be crucial to help support an evidence-based approach to global governance of disease.

## Data Availability Statement

Data are available in a public, open access repository [dataset] Ljungqvist GV, Weets CM, Stevens T, Robertson H, Zimmerman R, Graeden E, et al. Data from: Global Patterns in Access and Benefit-Sharing: A Comprehensive Review of National Policies [Internet]. GitHub; 2024. Available from: https://github.com/cghss/ABS

