## Supplementary Figure 1 for "Global Patterns in Access and Benefit-Sharing: A Comprehensive Review of National Policies": ABS Descriptive Supplemental Fig1.docx

### Supplementary Material

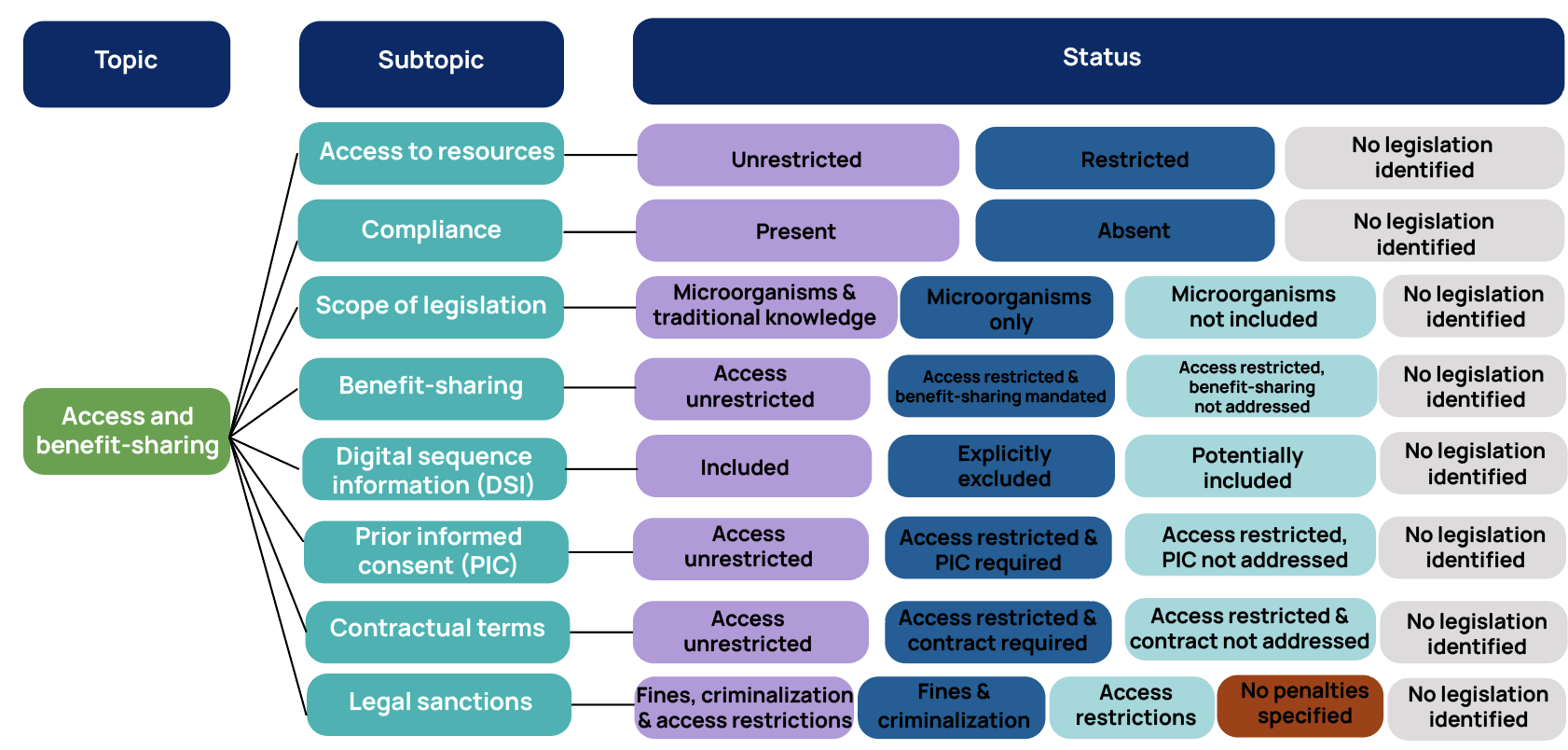

**Supplementary Figure 1: Data taxonomy used in Analysis and Mapping of Policies for Emerging Infectious Diseases Access and Benefits Sharing topic.** Each topic is split into subtopics, all of which ask a research question of the policy analyzed. Each of the UN Member States is then assigned an associated status for each of the subtopics based upon the policy identification, screening, analysis and verification processes.
