## Supplementary Table 1 for "Global Patterns in Access and Benefit-Sharing: A Comprehensive Review of National Policies": ABS Descriptive Supplemental Table 1.docx

### Supplementary Material

**Supplementary Table 1: Sequential Query Terms.** For each country, the following search terms were entered into Google, and all potentially relevant resources were reviewed. Any relevant policies identified in these resources or directly surfaced through the Google search were collected for further screening.

| Order | Query Term |
| --- | --- |
| 1 | **[Insert Country] “access benefit sharing”** |
| 2 | **[Insert Country] “digital sequence information”** |
| 3 | **[Insert Country] “genetic resource”** |
