## Supplementary Table 2 for "Global Patterns in Access and Benefit-Sharing: A Comprehensive Review of National Policies": ABS Descriptive Supplemental Table 2.docx

### Supplementary Material

**Supplementary Table 2: Inclusion criteria for policies included in each subtopic.** Inclusion criteria are developed through literature review and with input from subject matter experts. These criteria are applied to each policy during the screening and analysis processes to determine their applicability to the project and to each subtopic.

| Subtopic | Inclusion Criteria |
| --- | --- |
| Access to Resources | Included documents were analyzed for the restrictions they imposed on access to genetic resources. Policies that addressed restrictions on the access of international entities to domestic resources were included in this dataset. Legislation that explicitly stated that access to domestic resources was unrestricted, meaning that any foreign party could acquire and utilize genetic resources domestically free from fees or permits, were coded as unrestricted access. If any fees, permits, consent-seeking procedure, or benefit-sharing mechanism were incurred for access, this was coded as restricted. |
| Compliance | Compliance mechanisms were defined as language within national legislation empowering the government to ensure domestic resources were legally accessed. Here, countries that had ABS- relevant legislation without access restrictions were included, as many such countries still required compliance with foreign countries' legislation. Countries were coded as either having compliance mechanisms outlined or not. |
| Scope of legislation | Included documents were analyzed for the scope of their application. Applicable laws may have originally been drafted for various purposes, including biological conservation, biotechnology, and scientific research. Furthermore, some laws predated the widespread use of genetic technology. As a result, the terminology used in the law varied, and with it, its scope of application. Approaching laws from the perspective of pathogen-sharing, the research team found three broad interpretations of the scope described below:  The modern gold standard, as defined by the Nagoya Protocol, applied legislation to all genetic material and associated traditional knowledge. Another option was to establish the scope as genetic material alone, with no mention of traditional knowledge. A third option used a variety of specific scopes such as “flora and fauna”, “plants and animals”, or any other phrase which failed to capture micro-organisms such as bacteria and viruses. Countries that had legislation solely covering traditional knowledge without genetic material and legislation that was deemed too broad, such as covering “all natural resources”, were excluded. |
| Benefit-sharing | Benefit-sharing refers to the process of sharing any incurred benefits from the accessed genetic resource. This can take a monetary form (such as royalties, access fees, and milestone payments), or a non-monetary form (such as scientific collaboration, capacity-building, or technology transfer). Countries with unrestricted access were excluded from this subtopic, as this was de facto not applicable. Countries with access restrictions were divided according to whether they had outlined benefit-sharing requirements, or not. |
| Digital sequence information (DSI) | Laws that explicitly or conceivably pertain to Digital sequence information were included in this dataset. Due to the recent adoption of the term “DSI”, laws that used unclear terminology that the research team concluded could be interpreted to include DSI were included. Where supporting documents existed that clarified the country’s interpretation of nebulous laws, such documents were also uploaded. |
| Prior Informed consent (PIC) | PIC refers to a process whereby local government entities or local communities need to consent to the acquisition and/or utilization of the genetic resources priorly. In accordance with the Nagoya protocol, this must also be fully informed, meaning that they must be provided with the full information on volume, purpose, and time of access. Countries with unrestricted access were excluded from this, as this was de facto not applicable. Countries with restricted access were differentiated into whether they had an outlined prior consent requirement of any kind, or not. |
| Contractual terms | Many countries require a formalized contract-setting process to access genetic resources, often with terms relating to the agreed-upon benefits to be shared. This can take various forms, including Mutually Agreed Terms, Material Transfer Agreements, ABS contracts, and more. Some countries furthermore provide model contracts, or strict guidance, to facilitate the process. Countries with unrestricted access were excluded from this, as this was de facto not applicable. Countries with access restrictions were divided according to contract requirements of any kind, or not. |
| Legal sanctions | Legal sanctions refers to provisions in laws specifically applicable to the unauthorized access, export, and/or utilization of genetic resources. Here too, countries without access restrictions were included, as many such countries still required compliance with foreign countries' legislation. This was divided according to type, such as fines and criminalization (including incarceration), access restrictions (confiscations, revocations, or bans), or both. |
